# Sex-specific difference of long-term mortality after transcatheter edge-to-edge repair for functional mitral regurgitation; Insights from the OCEAN-Mitral Registry

**DOI:** 10.1101/2023.03.28.23287886

**Authors:** Hirofumi Hioki, Yusuke Watanabe, Akihisa Kataoka, Ken Kozuma, Shinichi Shirai, Toru Naganuma, Masahiro Yamawaki, Yusuke Enta, Shingo Mizuno, Hiroshi Ueno, Yohei Ohno, Yoshifumi Nakajima, Masaki Izumo, Hiroki Bouta, Kazuhisa Kodama, Junichi Yamaguchi, Shunsuke Kubo, Makoto Amaki, Masahiko Asami, Mike Saji, Kazuki Mizutani, Shinya Okazaki, Daisuke Hachinohe, Toshiaki Otsuka, Yuya Adachi, Masanori Yamamoto, Kentaro Hayashida, OCEAN-Mitral Investigators

**Author notes:** **Address for correspondence:** Hirofumi Hioki, MD, PhD, Division of Cardiology, Teikyo University School of Medicine, 2-11-1 Kaga, Itabashi-ku, Tokyo 173-8605, Japan.

## Abstract

**Background:** Recent studies suggested short-term mortality after transcatheter edge-to-edge repair (TEER) was comparable between male and female. However, the sex-specific prognostic difference in long-term follow-up after TEER is still unknown. To evaluate the impact of sex on long-term mortality after TEER for functional mitral regurgitation (FMR) using multicenter registry data.

**Methods:** We retrospectively analyzed 1220 patients (male 60.3%) who underwent TEER for FMR at 24 centers. Impact of sex on all-cause death and hospitalization for heart failure (HF) after TEER was evaluated using multivariate regression analysis and propensity score (PS) matching methods.

**Results:** During the two-year follow-up, 205 all-cause death and 259 hospitalizations for HF were observed after TEER for FMR. Male had a significantly lower incidence of all-cause death than female (18.7% vs. 14.0%, log-rank p < 0.01). After adjustment by multivariate Cox-regression and PS matching, male was significantly associated with a higher incidence of all-cause mortality after TEER than female (hazard ratio [HR] 2.24, 95% confidence interval [CI] 1.48 to 3.39 in multivariate Cox-regression; HR 2.04, 95% CI 1.17 to 3.57 in PS matching). The sex-specific prognostic difference was even more pronounced after 1-year of TEER. On contrary, there was no sex-related difference in hospitalization for HF after TEER.

**Conclusions:** Male had a higher incidence of two-year all-cause death during after TEER for FMR than female, while this was not observed in hospitalization for HF. This result might indicate that female with FMR is more likely to benefit from TEER for better prognosis.

**What is known?:** There were conflicting results in sex-related prognostic difference after TEER for FMR.

**What the study adds:** During two-year follow-up period, male had higher increased risk of mortality after TEER than female in multivariate regression and propensity score matched analysis, while there was no sex-related difference in hospitalization for HF after TEER. Female with FMR might be likely to derived benefit from TEER concomitant with GDMT as compared to male.

## Introduction

With the aging population, because of its high mortality and other adverse events, the increasing prevalence of functional mitral regurgitation (FMR) become a great concern [1,2]. Based on the recent clinical guidelines, transcatheter edge-to-edge repair (TEER) is recommended in selected population with FMR, particularly in high surgical risk [3,4].

Among the vast majority of cardiovascular diseases, sex difference has been described in prevalence, pathophysiology, clinical scenario, and outcomes [5]. In valvular heart disease, surgical valve replacement or repair is related to higher mortality and complications in female than in male [6,7]. Moreover, female, who outlive male, might be supposed to suffer from heart failure because aging is associated with heart failure progression [8]. Recently, the advancement of transcatheter valve therapy renewed sex-specific differences and the previous studies demonstrated better prognosis in female than male undergoing transcatheter aortic valve implantation [9]. Therefore, the question of whether sex-related prognostic differences still exist after TEER for FMR should be clarified.

While the previous registry data showed comparable prognosis between male and female after TEER, the recent meta-analysis shed light on the sex-related prognostic differences after TEER in that female had lower mortality on long-term follow-up than male [10,11]. Therefore, we sought to verify the impact of sex on long-term mortality after TEER for FMR using multicenter registry data.

## Methods

### Study population

This is a retrospective analysis of the Optimized transCathEter vAlvular interveNtion (OCEAN) transcatheter mitral valve intervention (OCEAN-Mitral) registry. The OCEAN-Mitral registry is a prospective, multicenter, observational registry to assess the real-world clinical outcomes of TEER at 24 collaborating hospitals located in Japan. This study was registered with the University Hospital Medical Information Network Clinical Trials Registry, as accepted by the International Committee of Medical Journal Editors (UMIN000023653).

The study protocol was developed in accordance with the Declaration of Helsinki and was approved by the ethics committee of each participating hospital. All patients gave informed consent before participating in this study. Between April 2018 and June 2021, 2150 patients underwent TEER at each center. After excluding TEER for primary MR, cases with missing data, and in-hospital death, a total of 1220 patients treated with TEER for FMR were included in this analysis (**Figure 1**). Baseline data including patients’ comorbidities, findings of echocardiography, and procedural details were collected. Echocardiographic assessment of MR severity and the procedure of TEER using MitraClip NT, NTW, XT, XTW (Abbott Structural Heart, Santa Clara, California) had performed at the discretion of the interdisciplinary heart team in each participating hospital.

**Figure 1.**
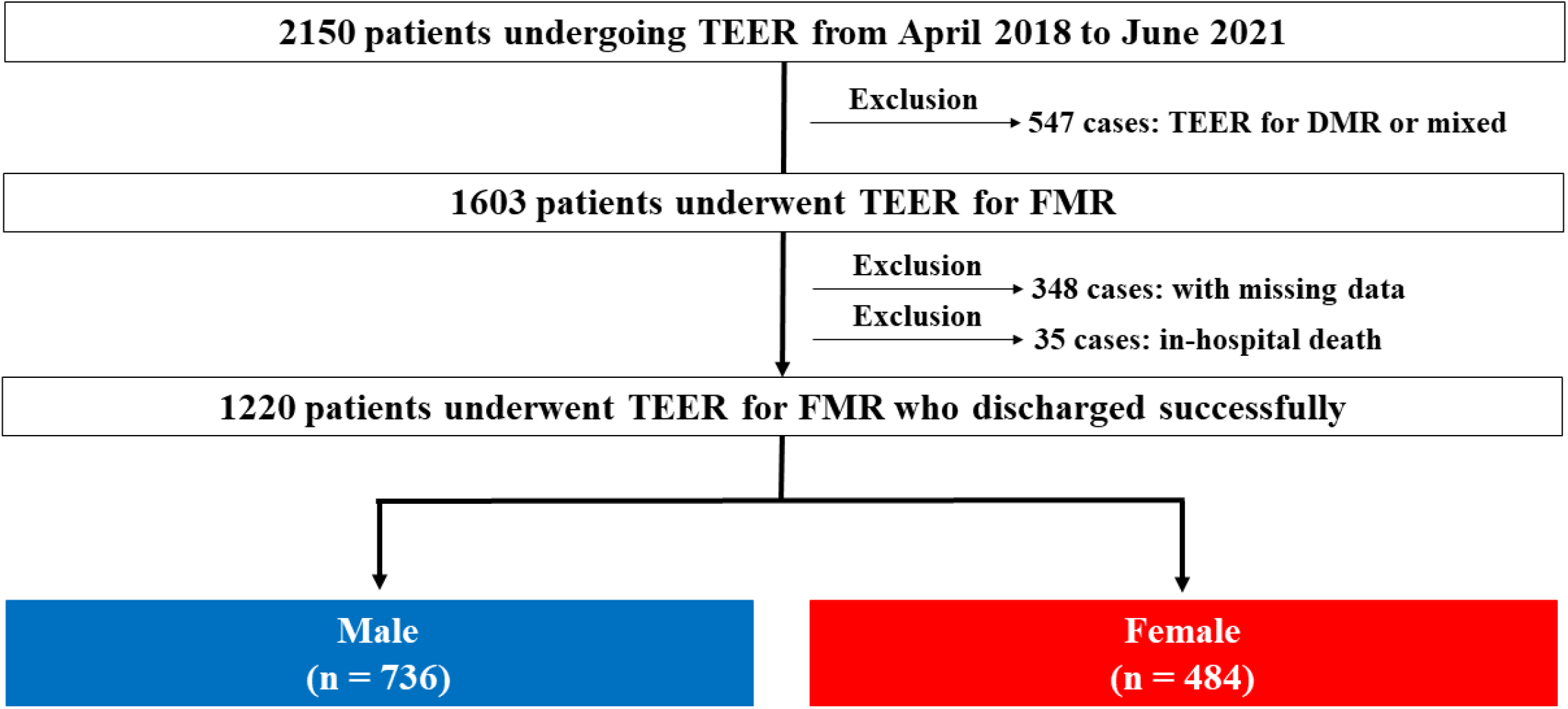
Study population. FMR, functional mitral regurgitation; MR, mitral regurgitation; TEER, transcatheter edge-to-edge repair.

### Clinical endpoint

The endpoints of this study were all-cause death and hospitalization for heart failure (HF) after TEER during two-year follow-up. Survival status was collected through either outpatient visit or telephone interview with patients, or patients’ relatives.

### Statistical analysis

Continuous variables were assessed for normal distribution using the Shapiro-Wilk test and presented as median (interquartile range [IQR]). Dichotomous variables were described as numbers and percentages. Based on the hypothesis that the effect of treatment, including medical therapy and/or TEER, might have mutual interaction with the organism of each sex, we divided all patients into two groups according to male and female. Differences between the two groups were compared using the chi-square test for categorical variables and Student’s t-tests or Wilcoxon rank-sum tests, as appropriate, for continuous variables. The Kaplan-Meier test was used to estimate the incidence of two-year all-cause death and hospitalization for HF after TEER and the difference in survival between male and female was compared using a log-rank test and Gray test. Multivariate Cox-regression analysis was applied to evaluate the impact of sex (being male) on two-year all-cause death after TEER as compared to that of female. The assumption of proportionality was examined using a log-minus-log plot for all-cause death according to sex. To estimate the impact of sex on hospitalization for HF, we used the Fine-Gray competing risk regression model to account for the competing risk of death and generated subdistribution hazard ratio (sub-HR). Confounders in multivariate analysis were determined based on the clinical significance and multicollinearity. To reduce the potential confounding effects due to background variability in the direct comparison between male and female, a propensity score (PS) for being male was estimated using logistic regression. The details of final multivariate model and the variables entered into PS calculation were described in Supplementary Methods. PS matching was performed using the nearest neighbor on the logit of the PS for being male with caliper width set to 0.2. The validity of the model to generate covariate balance between matched groups was evaluated using standardized mean differences on whether an appropriate balance was secured across PS-matched groups [12]. Statistical analysis was performed using the R software (Version 3.0.2, The R Project) and the Statistical Package for Social Science, version 21 (SPSS Inc., Chicago, IL, USA) software. A p-value was 2-sided and a value < 0.05 was considered statistically significant.

## Results

### Baseline patients’ characteristics

Of the 1220 patients, 736 patients (60.3%) were male, while the remaining 484 patients (39.7%) were female. Baseline patient characteristics of the study population are shown in Table 1. There was significant difference in baseline characteristics between the two groups, except for the presence of NYHA functional class III/IV, hypertension, atrial fibrillation, chronic kidney disease, beta blocker, mineralocorticoid receptor antagonist, angiotensin receptor neprilysin inhibitor, and SGLT-2. Echocardiographic findings before TEER are shown in Table 2. Compared to male, female had smaller left ventricular end-diastolic volume index and left atrial diameters. Female also had lower proportion of ventricular FMR with ischemic cardiomyopathy, lower regurgitation volume, and lower pulmonary artery systolic pressure as compared to male.

**Table 1.** Baseline patient characteristics.

|  | Male | Female |  |
| --- | --- | --- | --- |
| Variable | (n = 736) | (n = 484) | p-value |
| Age (years) | 77 (70-83) | 80 (74-85) | <0.01 |
| Body surface area (/m<sup>2</sup>) | 1.62 (1.51-1.73) | 1.38 (1.29-1.47) | <0.01 |
| New York Heart Association III or IV | 467 (63.5%) | 327 (67.6%) | 0.16 |
| Frail (Clinical frailty scale ≥5) | 138 (18.8%) | 156 (32.2%) | <0.01 |
| Smoking | 399 (54.3%) | 62 (12.8%) | <0.01 |
| Hypertension | 511 (69.4%) | 316 (65.3%) | 0.13 |
| Diabetes Mellitus | 265 (36.0%) | 128 (26.4%) | <0.01 |
| Dyslipidemia | 440 (59.8%) | 235 (48.6%) | <0.01 |
| Atrial fibrillation | 491 (66.7%) | 319 (65.9%) | 0.80 |
| Chronic kidney disease | 631 (85.7%) | 433 (89.5%) | 0.07 |
| Coronary artery disease | 348 (47.3%) | 152 (31.4%) | <0.01 |
| Coronary revascularization | 319 (43.3%) | 136 (28.1%) | <0.01 |
| Stroke | 101 (13.7%) | 40 (8.3%) | <0.01 |
| Any implantable pacing device | 215 (29.2%) | 109 (22.5%) | 0.01 |
| Cardiac resynchronization therapy | 113 (15.4%) | 47 (9.7%) | <0.01 |
| History of open-heart surgery | 103 (14.0%) | 51 (10.5%) | 0.08 |
| Chronic obstructive pulmonary disease | 87 (11.8%) | 28 (5.8%) | <0.01 |
| STS risk score (Mitral valve replacement) | 8.86 (5.27-13.73) | 9.88 (6.38-14.94) | <0.01 |
| Medication before transcatheter mitral valve repair |  |  |  |
| Angiotensin-converting enzyme inhibitor<br>or angiotensin receptor blocker | 411 (55.8%) | 250 (51.7%) | 0.15 |
| Beta blocker | 611 (83.0%) | 400 (82.6%) | 0.88 |
| Mineralocorticoid receptor antagonist | 417 (56.7%) | 284 (58.7%) | 0.52 |
| Sodium-glucose cotransporter-2 inhibitors | 99 (13.5%) | 48 (9.9%) | 0.07 |
| angiotensin receptor neprilysin inhibitor | 14 (1.9%) | 9 (1.9%) | 1.00 |
| Loop diuretics | 635 (86.3%) | 423 (87.4%) | 0.61 |

Data are presented as median (25–75 percentiles), or numbers (percentages).

**Table 2.** Preprocedural echocardiographic finding.

|  | Male | Female |  |
| --- | --- | --- | --- |
| Variable | (n = 736) | (n = 484) | p-value |
| Left ventricle |  |  |  |
| Left ventricular diastolic diameter, mm | 62.0 (56.0-69.0) | 54.0 (48.0-61.0) | <0.01 |
| Left ventricular systolic diameter, mm | 52.0 (42.0-60.0) | 42.0 (34.0-52.0) | <0.01 |
| Left ventricular end-diastolic volume index, mL/m<sup>2</sup> | 108.2 (82.7-140.0) | 84.3 (60.3-118.2) | <0.01 |
| Left ventricular end-systolic volume index, mL/m<sup>2</sup> | 69.2 (44.1-96.4) | 48.1 (28.5-78.2) | <0.01 |
| Left ventricular ejection fraction, % | 35.0 (28.0-46.0) | 40.0 (31.0-55.0) | <0.01 |
| Mitral valve |  |  |  |
| Etiology of mitral regurgitation |  |  | <0.01 |
| Ventricular FMR with ischemic cardiomyopathy | 255 (34.6%) | 92 (19.0%) |  |
| Ventricular FMR without ischemic cardiomyopathy | 339 (46.1%) | 223 (46.1%) |  |
| Atrial FMR | 142 (19.3%) | 169 (34.9%) |  |
| Mitral regurgitation severity |  |  | 0.01 |
| Grade 2+ | 74 (10.1%) | 69 (14.3%) |  |
| Grade 3+ | 224 (30.4%) | 164 (33.9%) |  |
| Grade 4+ | 438 (59.5%) | 251 (51.9%) |  |
| Effective regurgitation orifice area, cm<sup>2</sup> | 0.33 (0.25-0.44) | 0.30 (0.21-0.38) | <0.01 |
| Regurgitation volume, ml | 50.0 (39.3-65.0) | 46.0 (34.8-60.0) | <0.01 |
| Left atrial diameter, mm | 51.0 (46.0-58.0) | 48.0 (42.0-54.0) | <0.01 |
| Pulmonary artery systolic pressure, mmHg | 41.0 (31.0-53.0) | 36.0 (28.0-49.0) | <0.01 |

Data are presented as median (25–75 percentiles), or numbers (percentages).

### Procedural outcomes

Although procedural time and number of clips during procedure were different between male and female, the achievement of MR reduction during procedure was comparable between two groups (Table 3). Post-procedural transmitral pressure gradient was higher in female than male.

**Table 3.** Procedural index and postprocedural echocardiographic findings.

| Variable | Male<br>(n = 736) | Female<br>(n = 484) | p-value |
| --- | --- | --- | --- |
| Procedure time, minutes | 90.0 (64.0-122.0) | 83.5 (62.0-111.8) | 0.01 |
| Anesthesia time, minutes | 154.0 (124.0-200.0) | 144.0 (115.0-174.0) | <0.01 |
| Number of Clip during procedure |  |  | <0.01 |
| 1 | 427 (58.0%) | 339 (70.0%) |  |
| 2 | 293 (39.8%) | 142 (29.3%) |  |
| 3 | 16 (2.2%) | 3 (0.6%) |  |
| Mitral regurgitation severity |  |  | 0.68 |
| Grade 1+ | 571 (77.6%) | 377 (77.9%) |  |
| Grade 2+ | 144 (19.6%) | 93 (19.2%) |  |
| Grade 3+ | 14 (1.9%) | 12 (2.5%) |  |
| Grade 4+ | 7 (1.0%) | 2 (0.4%) |  |
| Improvement of mitral regurgitation $\geq 1$ grade | 735 (99.9%) | 484 (100.0%) | 1.00 |
| Transmitral mean pressure gradient, mmHg | 2.5 (2.0-3.5) | 3.0 (2.0-4.0) | <0.01 |

Data are presented as median (25–75 percentiles), or numbers (percentages).

### Clinical outcomes

The median follow-up duration was 445 (IQR, 364 to 729) days. During the follow-up period, there were 205 cases (16.8%) of all-cause death, including 139 cases (11.4%) of cardiovascular (CV) death, and 259 cases (21.2%) of hospitalization for HF. Incidence of two-year all-cause death was significantly higher in male than female, and moreover, it was mainly driven by relatively high incidence of CV death during two-year follow-up (**Figure 2**).

**Figure 2.**
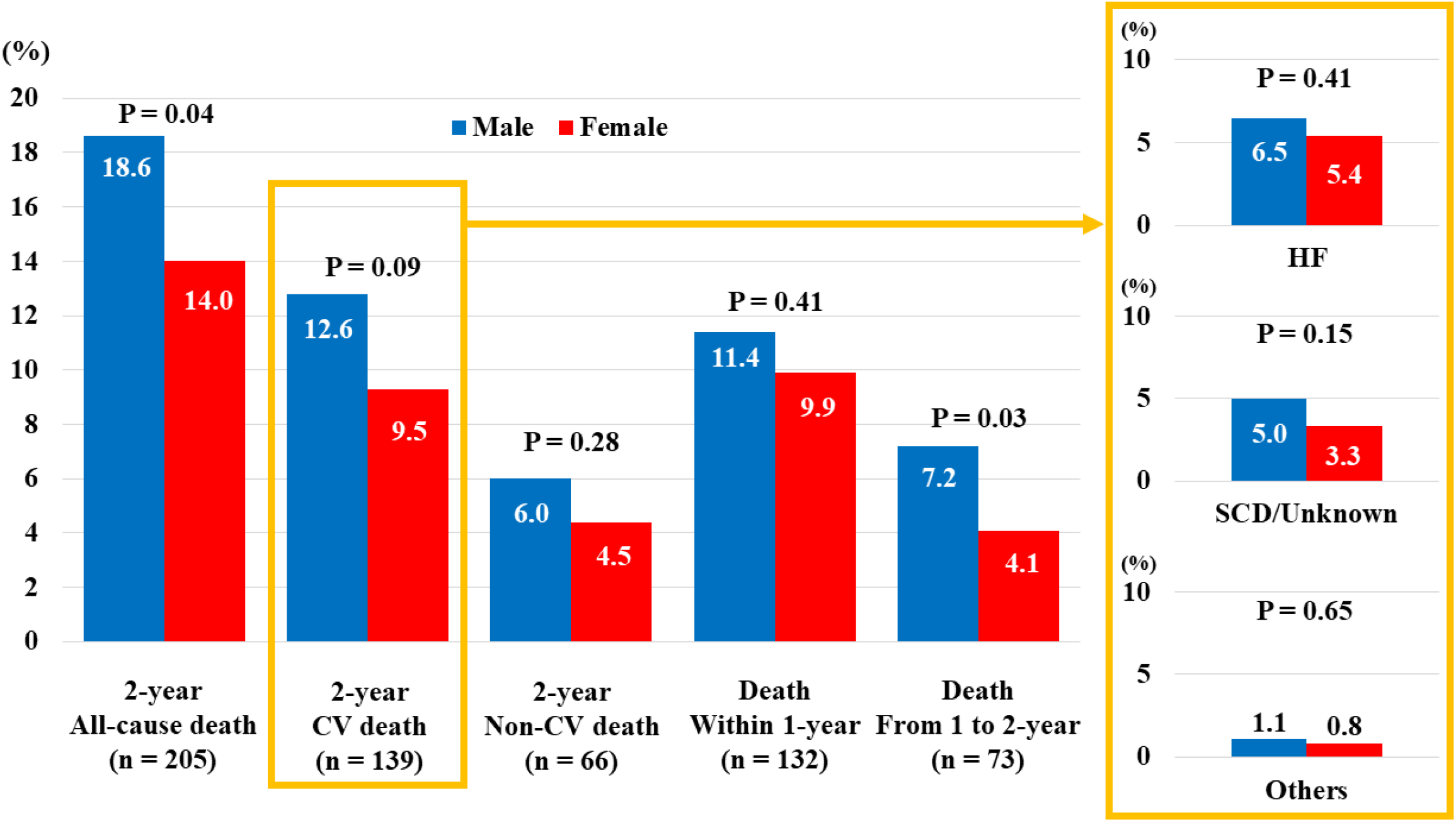
Comparison of survival during two-year follow-up after TEER between male and female. Male had higher incidence of two-year all-cause and cardiovascular (CV) death during the follow-up. Sex-related prognostic difference was more pronounced after one-year of TEER. HF, heart failure; SCD, sudden cardiac death.

The details of two-year CV death included 74 cases of death due to HF (male vs. female, 6.5% vs. 5.4%, p = 0.41), sudden cardiac death or unknown cause (male vs. female, 5.0% vs. 3.3%, p = 0.15), and other causes (male vs. female, 1.1% vs. 0.8%, p = 0.65). The Kaplan-Meier curve demonstrated that the incidence of all-cause death was significantly higher in male than female (18.7% vs. 14.0%, log-rank P = 0.03) (**Figure 3A**). Multivariate Cox-regression analysis revealed that male had an increased risk of two-year all-cause death as compared to female (adjusted hazard ratio[HR] 2.24, 95% confidence interval [CI] 1.48 to 3.39, p < 0.01). PS matching was performed to characterize the survival outcomes in population with matched baseline characteristics. Patients’ baseline characteristics in the matched cohort were relatively well balanced between male and female (Table S1). In the PS matched cohorts, male had also higher increased risk of all-cause death than female (HR 2.04, 95%CI 1.17 to 3.57, p = 0.01) (**Figure 3B**). On contrary, the incidence of hospitalization for HF after TEER was comparable between male and female in multivariate analysis (**Figure 4A**) and PS matching analysis (**Figure 4B**). The multivariate competing risk regression analysis confirmed that male was not significantly associated with an increased risk of hospitalization for HF as compared to female in overall (adjusted sub-HR 1.43, 95%CI 0.99 to 2.08, p = 0.06). This result was consistent in the competing risk regression analysis in the matched cohort (HR 1.34, 95%CI 0.81 to 2.21, p = 0.25).

**Figure 3.**
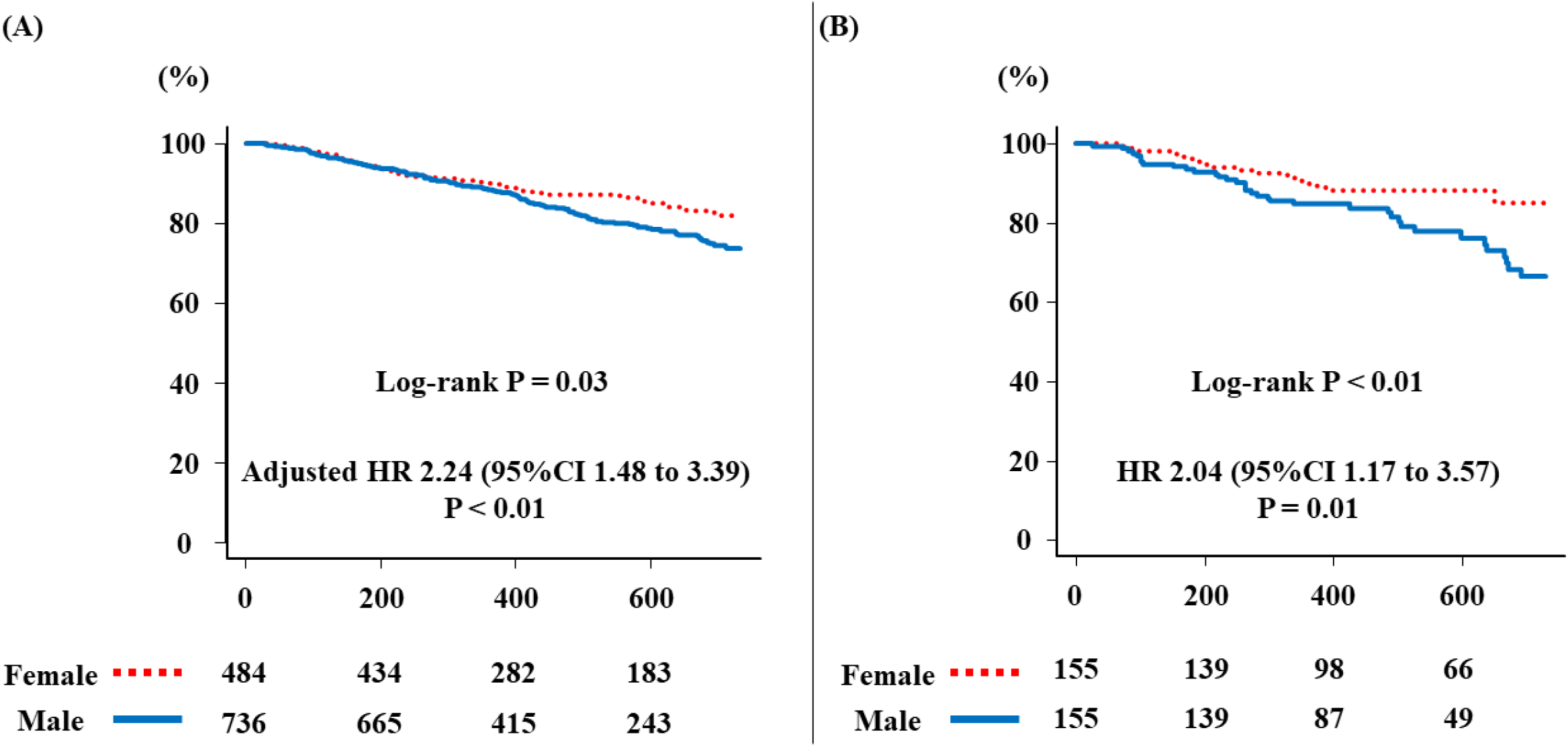
The Kaplan-Meier analysis for freedom from two-year all-cause death after TEER. **(A)** Male had higher incidence of all-cause death than female (18.6% vs. 14.0%, log-rank P = 0.03). **(B)** The result was consistent with analysis in the matched cohorts (log-rank P < 0.01).

**Figure 4.**
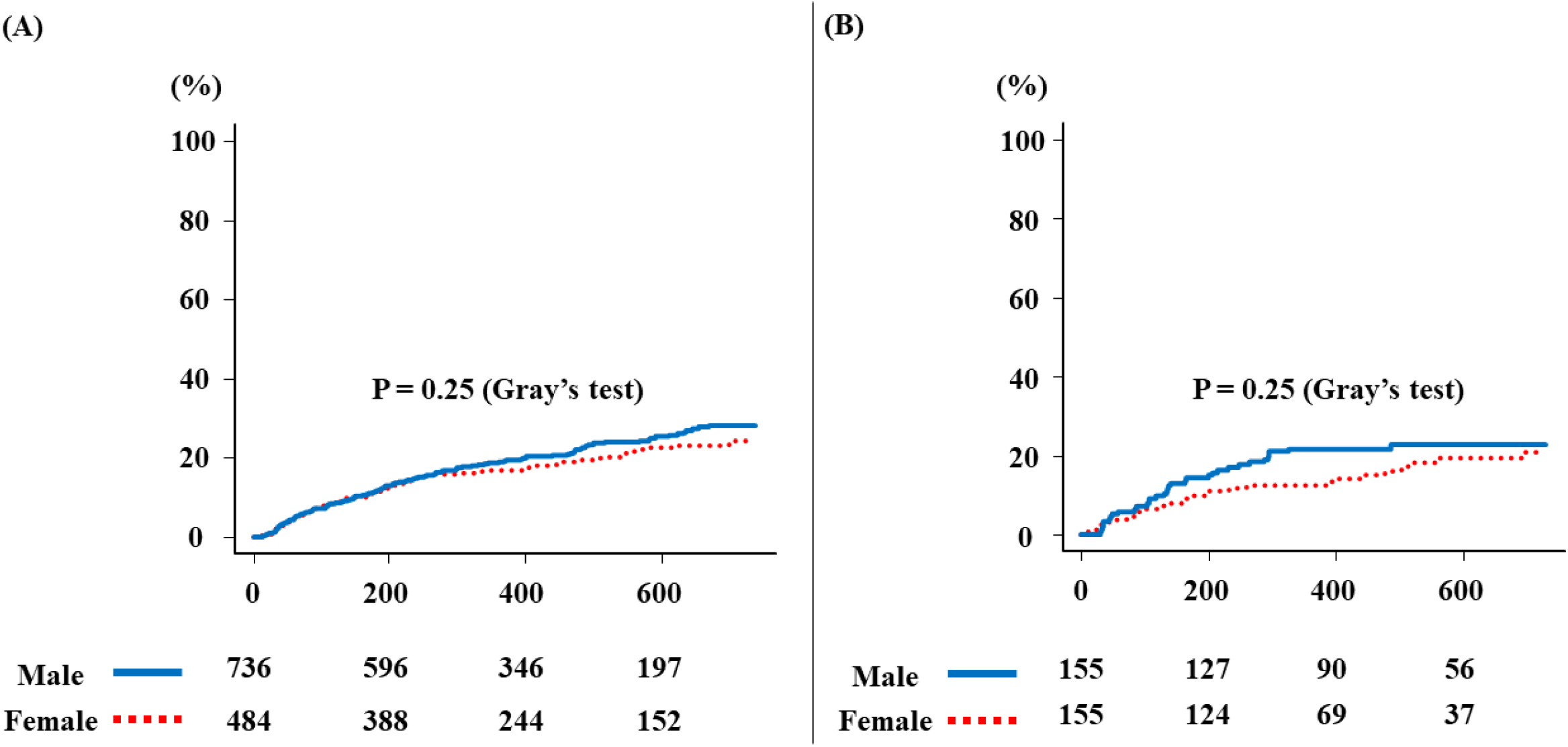
The Kaplan-Meier analysis for hospitalization for heart failure after TEER. **(A), (B)** The incidence of hospitalization for HF were comparable between male and female in all cohort and the matched cohorts.

## Discussion

In the current analysis using the multicenter registry data, sex-related differences in patients’ characteristics and clinical outcomes were evaluated between male and female who underwent TEER using MitraClip for FMR. The main findings of this analysis are as follows: 1) female had higher age, smaller left ventricular volume, and smaller EROA as compared to male, whereas male displayed greater prevalence for the comorbidities (e.g., diabetes, CAD, pacing device, or COPD) than female; 2) male had significantly higher incidence of all-cause death after TEER for FMR, as compared to female, 3) the sex specific prognostic difference tended to increase gradually after one-year of TEER, and 4) the incidence of hospitalization for HF after TEER was comparable between male and female. To our knowledge, this is the first report demonstrating the impact of sex on long-term mortality after TEER, and moreover, there was a trend to increase sex-specific difference on mortality after TEER after one-year of the procedure.

Although surgical mitral valve repair has been a conventional treatment for patients with FMR, the prognosis of female was reported as worse than male after surgical mitral valve repair for FMR [6,13]. Because the current guideline recommended the same cutoff value to quantify MR severity in both male and female, the severity of MR in female is likely to be underestimated and this resulted in delayed mitral valve intervention since female usually have smaller left ventricular anatomy and higher LVEF [3,4]. In aortic stenosis, likewise, perioperative mortality and complications are higher in female than male who undergo surgical aortic valve replacement, however, transcatheter aortic valve therapy has succeeded to reduce the sex-related prognostic difference and mid-term survival is greater in female [14]. According to these findings, the incidence of mortality after TEER is presumed to be lower in female, though the previous studies have failed to reach consistent results due to heterogeneity of baseline characteristics [10,15]. For this reason, the current analysis sought to evaluate the sex difference in mortality after TEER using crude and matched cohorts of multicenter registry data.

Historically, male had a higher incidence of CV death than female, particularly at the period in life [16]. The current analysis corroborated in that male had higher all-cause death, which was mainly due to a higher incidence of CV death, after TEER for FMR than female. Recently, the COAPT risk score, composed of comorbidities and echocardiographic parameters, dedicated to provide useful risk stratification in patients undergoing TEER [17]. Because the prevalence of COPD, LV dysfunction, and large LV volume, which are components of the COAPT risk score, were greater in male than female in our cohort, it might be a reason why female had a lower incidence of all-cause death than male. Because patients’ baseline characteristics in our cohort were consistent with those of the EuroSMR registry, our population might reflect the real-world population undergoing TEER [10]. However, even though the patients’ characteristics were similar between the current analysis and the EuroSMR registry, the clinical result, particularly mortality, was completely different. This difference might be related to the different proportion of etiology of FMR, demonstrating that female in the current analysis had low VFMR with ICM and high atrial FMR whereas nearly half of female in the EuroSMR registry had VFMR with ICM. Based on the previous data of worse prognosis of VFMR as compared to atrial FMR, female in the current study had relatively better prognosis after TEER than those of other study [18]. and therefore, the prognostic. As the authors of EuroSMR registry mentioned unmeasured confounding variables in their limitation, and therefore, we performed adjustments using the PS method in addition to the conventional multivariate regression analysis.

An interesting finding of our analysis was that the sex-related benefit of TEER on survival was more prominent after 1-year of the procedure and the incidence of CV death after 1-year of TEER tended to be high in male as compared to female. The recent meta-analysis including 24905 patients undergoing TEER also demonstrated that female had a significantly lower incidence of all-cause death during long-term follow-up, while there was no difference in survival between male and female during short-term (up to 30-days of TEER) [11]. Among the predictor of long-term survival after TEER, the recent study demonstrated that residual MR severity ≤ 1+ and absence of left ventricular dilatation were associated with better clinical survival, and indeed, nearly 80% of both male and female obtained residual MR severity ≤ 1+ and male had higher proportion of left ventricular dilatation in our cohort [19]. Moreover, the previous study evaluating sex-related prognostic differences in patients with advanced heart failure showed that female had a lower risk of mortality than male in heart failure due to non-ischemic heart disease, which accounted for more than half of FMR in this cohort [20]. These might be reason why the trend of sex-related prognostic difference was even more pronounced after one-year of TEER in the current analysis. Because CV death was the main cause of death after TEER, an intensive titration of guideline-direct medical therapy is advocated to achieve better prognosis in both male and female.

We also found that the reduction of hospitalization for HF after TEER was comparable between male and female. Though TEER similarly improves severity of FMR in both male and female, the COAPT sub-analysis demonstrated that the effectiveness of TEER in reducing HF was diminished after the first year in female as compared to male [21]. On contrary, the response to pharmacological therapy, including beta blockers, and CRT were better in female than male [22,23]. In the current cohort, the proportion of beta-blocker and other drugs were comparable between male and female, and therefore, based on the previous studies, female in our cohort were likely to be derived benefit from drugs of HF even when the effectiveness of TEER to reduce HF after the first year as compared to male. These might be reason why our population had neutral effect on suppression of hospitalization for HF after TEER.

Eventually, the current analysis demonstrated the sex-related prognostic difference after TEER for FMR and the prognostic difference was even more pronounced after one-year of TEER. As transcatheter valve intervention improved survival in female with severe aortic stenosis as compared to surgical valve replacement, we believed that female with FMR would be more likely to benefit from TEER for better clinical outcomes.

## Limitations

There were several limitations in this study. First, this is the retrospective analysis of prospective multicenter registry data. Therefore, there were several confounding variables between male and female. To reduce the bias and confounding, the current analysis performed the PS matching analysis in addition to the conventional multivariate regression analysis.

Second, the impact of medications on survival after one-year of TEER could not be fully evaluated because the data concerning titration of drugs for heart failure after TEER were not collected through the follow-up duration, though the proportion of drugs for heart failure at baseline in the current analysis were almost similar to those of the COAPT trial [24]. Third, the echocardiographic parameters were not evaluated in an independent core laboratory. And fourth, 23.9% of patients with FMR were excluded from the final analysis due to missing data of pre-procedural echocardiographic parameters and in-hospital death. However, because the baseline patients’ characteristics between included and excluded patients were comparable (Table S2), our result would not be substantially changed if the excluded population were enrolled into the analysis.

## Conclusion

The current analysis demonstrated that male had higher incidence of all-cause death after TEER than female and sex-related prognostic difference was even more prominent after 1-year of TEER. Further study might be required to evaluate the clinical efficacy of sex-specific criteria of TEER for FMR to establish appropriate management strategies.

## Data Availability

The data underlying this article cannot be shared publicly due to the privacy of individuals that participated in the study. The data will be shared on reasonable request to the corresponding author.

## Acknowledgement

The authors thank all investigators participating in this registry.

## Supplementary Material

Supplementary Methods

Tables S1–S7

## Abbreviations List

FMR: functional mitral regurgitation
HF: heart failure
TEER: transcatheter edge-to-edge repair

